# Outcomes of primary percutaneous coronary intervention in a population of Iranian patients with myocardial infarction, according to the history of diabetes from 2018 till 2021

**DOI:** 10.1101/2025.10.27.25338920

**Authors:** Ensieh Sadat Mansouri, Sareh Niliahmadabadi, Abbas Soleimani, Peimaneh Heydarian, Zahra Shajari, Mostafa Rouzitalab, Shahrokh Karbalai Saleh

## Abstract

Primary percutaneous coronary intervention (PPCI) is the mainstay treatment for patients with ST-Elevation Myocardial Infarction (STEMI). However, depending on patient characteristics, this procedure may be associated with major adverse cardiovascular events (MACE). Diabetes mellitus has consistently been identified as a significant risk factor for adverse outcomes. In this study, we also evaluated the impact of other clinical factors; including hypertension, chronic kidney disease (CKD) and past medical history on MACE. In this retrospective cohort study, 722 patients who underwent PPCI at Sina Hospital between 2018 and 2021 were enrolled. Participants were categorized into two equal groups: 361 with diabetes and 361 without. Demographic information, PPCI-related characteristics, and clinical variables such as blood pressure, lipid profile, and CKD status were collected. Major outcomes, including myocardial infarction (MI), cerebrovascular accident (CVA), and mortality, were assessed in both the short term (within 24 hours post-PPCI) and long term (six months post-PPCI). Diabetic patients were significantly older (mean age 61.5 vs. 57.93 years) and had a higher proportion of females. They also had a greater history of MI and prior PPCI. Overall mortality after PPCI was significantly higher in diabetic patients, smokers and those with both diabetes and hypertension. Long-term mortality was notably associated with the presence of CKD. Lower admission blood glucose levels were linked to increased CVA incidence, while recurrent MI was more common among patients with reduced ejection fraction. Age was positively correlated with the occurrence of MACE.

## 1 Introduction

Cardiovascular disorders (CVD) remain the most prevalent diseases worldwide [1, 2]. The mortality rate among diabetic patients with CVD is 2 to 5 times higher than in non-diabetic individuals [3] and more than half of all diabetic patients ultimately die due to coronary artery disease [4]. Hypertension, another common comorbidity, is also a well-established risk factor for CVD and is known to worsen the prognosis of myocardial ischemia. Over recent decades, therapeutic advancements such as lipid-lowering medications, PPCI and coronary artery bypass grafting (CABG) have markedly improved survival in patients with CVD. In 1990, the utilization rates of CABG and PPCI were roughly equivalent; however, PPCI has since experienced significant growth in clinical use [5]. Outcomes and complications following PPCI are influenced by a range of factors, including patient characteristics, procedural techniques, operator expertise and institutional capabilities [6]. Despite the introduction of modern therapeutic strategies, such as novel antiplatelet agents and drug-eluting stents, diabetic patients continue to experience worse outcomes following PPCI compared to non-diabetics (4). Diabetic kidney disease, which affects approximately 40%of diabetic individuals, independently increases the risk of MACE after PPCI. One study reported total mortality rates in diabetic patients post-PPCI as 3.02% in-hospital, 4.12% short-term, and 9.24% long-term, compared to 1.59%, 2.46% and 5.35% respectively in non-diabetic patients [7]. Several studies have identified diabetes as a contributor to increase in-hospital mortality, stent thrombosis, impaired circulation, distal embolization and other post-PPCI complications [8–11]. However, some investigations suggest no significant differences in short-term or major complications between diabetic and non-diabetic patients, and they do not support diabetes as an independent risk factor for in-hospital adverse clinical events following PPCI [12, 13]. Other evidence indicates that the coexistence of diabetes and hypertension is a stronger predictor of MACE following PPCI than either condition alone [14, 15]. Given the high prevalence of both diabetes and CVD in the Iranian adult population [1, 16], and the conflicting findings in existing literature, this study aims to evaluate the impact of diabetes and other clinical factors such as hypertension and CKD on PPCI outcomes. A better understanding of these associations is essential for reducing the clinical and social burden of CVD.

## 2 Methods

This retrospective cohort study was approved by the Research Ethics Committee of Tehran University of Medical Sciences. A total of 722 patients who underwent PPCI following STEMI at Sina Hospital, affiliated with Tehran University of Medical Sciences, between 2018 and 2021 were included in the study. Patients were stratified into two groups: 361 with diabetes and 361 without diabetes. Demographic data and PPCI-related characteristics were collected for all participants. Additional clinical variables, including blood pressure, lipid profile, and history of CKD, were also assessed. Major outcomes evaluated included MI, CVA, revascularization, and mortality, both in the short term (within 24 hours post-PPCI) and the long term (six months post-PPCI). Minor outcomes such as pulmonary edema and elevated serum creatinine levels were also analysed. Patient data were obtained from hospital medical records. Long-term follow-up was conducted through review of medical documents and telephone interviews. Patients with incomplete medical records or those who could not be reached by phone were excluded from the study.

## 3 Definition

Diabetes mellitus was confirmed if one or more of the following criteria were met: (1) treatment with insulin or an oral hypoglycemic agent or both; (2) fasting blood glucose levels ≥ 126 mg/dl and casual blood glucose levels of 200 mg/dl or over; and (3) either fasting or casual blood sugar levels higher than the above values and hemoglobin A1c levels of 6.5% or higher. Hyperlipidemia was confirmed if one or more of the following criteria were met: (1) treatment with lipid-lowering agents; (2) cholesterol level ≥ 200 mg/dl; and (3) triglyceride concentration ≥ 150 mg/dl. Hypertension was confirmed if one or more of the following were met: (1) treatment with antihypertensive medications; (2) systolic blood pressure of ≥ 130 mm Hg or diastolic blood pressure of ≥ 80 mm Hg or both. Acute kidney injury was confirmed in case of increase in serum creatinine ≥ 0.3 mg/dl within 48 hours after PPCI.

## 4 Analysis

The data was entered into the Statistical Package for Social Science (SPSS) for Windows, version 21. All quantitative variables are expressed as mean ± standard deviation and interpreted with t-test. Categorical variables were compared by Chi-square test. Statistical significance was assumed at a p-value ≤ 0.05.

## 5 Results

Diabetic patients were significantly older (mean age of 61.59 vs. 57.93, *p* < 0.001) and had a higher proportion of women (23.2% vs. 12.6%, *p* < 0.001). They also had a higher history of PCI (19.6% vs. 11.3%, *p* = 0.005) and MI (21.8% vs. 14.0%, *p* = 0.015).

**Table 1:** Demographic characteristic of the study population.

| Variable | No Diabetes | Diabetes | Total | P-value |
| --- | --- | --- | --- | --- |
| N | 486 (71.5%) | 194 (28.5%) | 680 (100.0%) |  |
| Age, mean (SD) | 57.93 (11.23) | 61.59 (10.54) | 58.97 (11.16) | < 0.001 |
| Sex |  |  |  |  |
| Female | 61 (12.6%) | 45 (23.2%) | 106 (15.6%) | < 0.001 |
| Male | 425 (87.4%) | 149 (76.8%) | 574 (84.4%) |  |
| History of PCI |  |  |  |  |
| No | 431 (88.7%) | 156 (80.4%) | 587 (86.3%) | 0.005 |
| Yes | 55 (11.3%) | 38 (19.6%) | 93 (13.7%) |  |
| History of MI |  |  |  |  |
| No | 386 (86.0%) | 151 (78.2%) | 537 (83.6%) | 0.015 |
| Yes | 63 (14.0%) | 42 (21.8%) | 105 (16.4%) |  |
| History of CABG |  |  |  |  |
| No | 437 (97.3%) | 184 (95.3%) | 621 (96.7%) | 0.194 |
| Yes | 12 (2.7%) | 9 (4.7%) | 21 (3.3%) |  |
| History of CVA |  |  |  |  |
| No | 442 (98.4%) | 189 (97.9%) | 631 (98.3%) | 0.646 |
| Yes | 7 (1.6%) | 4 (2.1%) | 11 (1.7%) |  |
| History of Heart Failure |  |  |  |  |
| No | 359 (80.0%) | 167 (86.5%) | 526 (81.9%) | 0.047 |
| Yes | 90 (20.0%) | 26 (13.5%) | 116 (18.1%) |  |

**Note:** N number of patients, hPCI history of primary cutaneous interventions, hMI history of MI, hCABG history of CABG, hCVA history of CVA, hHeart failure history of heart failure.

**Table 2:** Number of involved vessels and culprit vessel according to diabetes status.

| Variable | No Diabetes | Diabetes | Total | P-value |
| --- | --- | --- | --- | --- |
| Number of involved vessels |  |  |  |  |
| - | 10 (2.1%) | 6 (3.1%) | 16 (2.4%) | 0.023 |
| 2VD | 139 (28.7%) | 60 (30.9%) | 199 (29.3%) |  |
| 3VD | 156 (32.2%) | 79 (40.7%) | 235 (34.6%) |  |
| SVD | 180 (37.1%) | 49 (25.3%) | 229 (33.7%) |  |
| SVG | 2 (1.0%) | 2 (2.4%) | 4 (1.4%) |  |
| Culprit vessel |  |  |  |  |
| Diagonal artery | 2 (1.0%) | 3 (3.5%) | 5 (1.7%) | 0.219 |
| LAD | 95 (45.7%) | 41 (48.2%) | 136 (46.4%) |  |
| LMCA | 1 (0.5%) | 0 (0.0%) | 1 (0.3%) |  |
| LCX | 18 (8.7%) | 10 (11.8%) | 28 (9.6%) |  |
| Marginal artery | 4 (1.9%) | 4 (4.7%) | 8 (2.7%) |  |
| RCA | 86 (41.3%) | 25 (29.4%) | 111 (37.9%) |  |

**Note:** 2VD two-vessel disease, 3VD three-vessel disease, SVD small vessel disease, SVG saphenous vein disease, LAD left anterior descending artery, LMCA left main coronary artery, LCX left circumflex artery, RCA right coronary artery.

Total mortality following PPCI was significantly higher among diabetic patients, smokers, and individuals with combined diabetes and hypertension or diabetes with hypertriglyceridemia (*p* < 0.05). Higher post-procedural ejection fraction (coefficient < 1, *p* < 0.005) and elevated systolic blood pressure at admission (coefficient < 1, *p* < 0.005) were identified as protective factors against mortality. Short-term mortality was significantly higher in diabetic patients, smokers, and those with either diabetes and hypertension or diabetes with hyperlipidemia (*p* < 0.03). Long-term mortality was significantly associated with the presence of kidney disease (*p* = 0.044). Ejection fraction (coefficient 0.96, *p* = 0.006) was the only factor linked to out-of-hospital mortality.

**Table 3:**
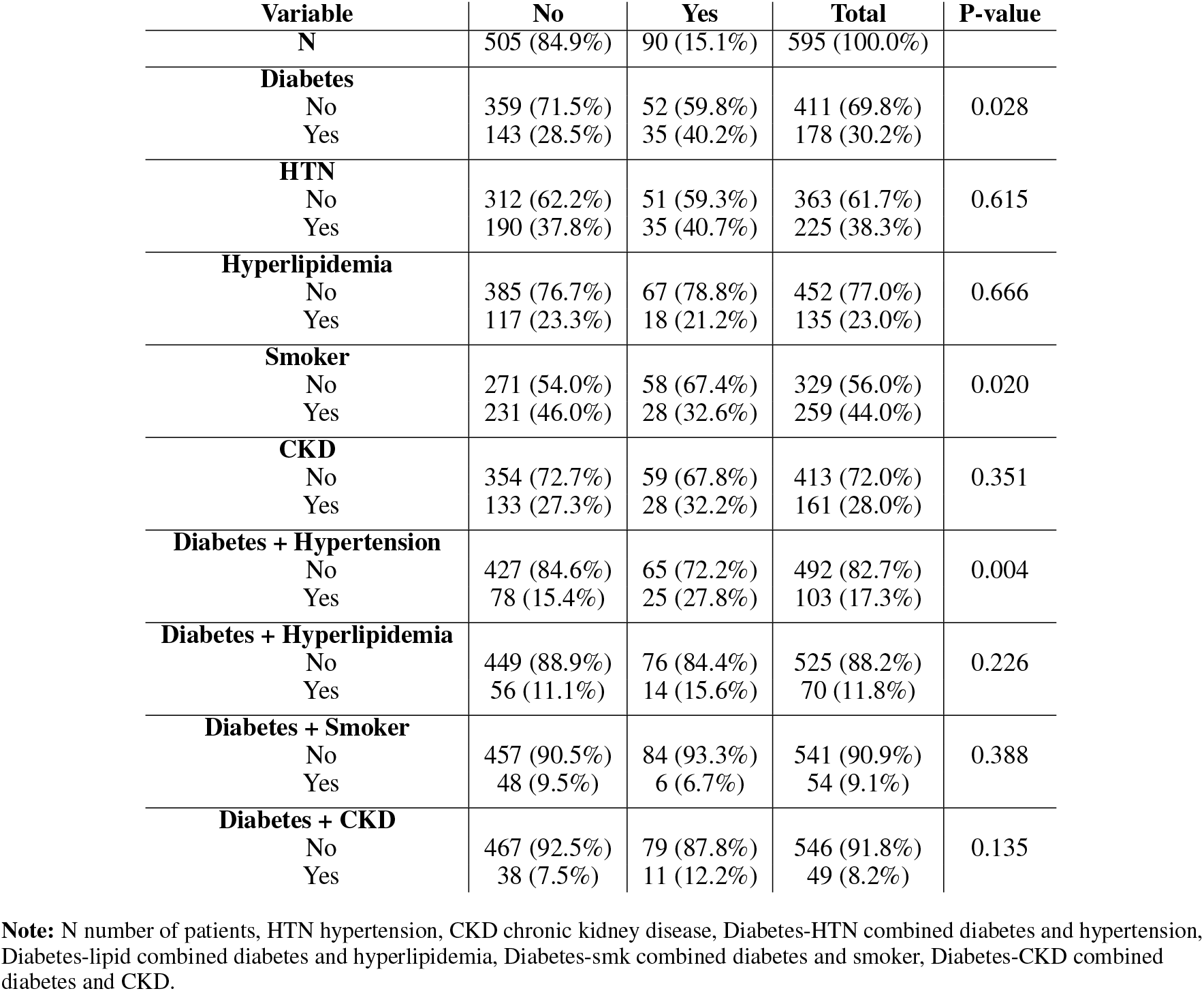
Mortality rate analysis in diabetics and non-diabetics.

**Table 4:**
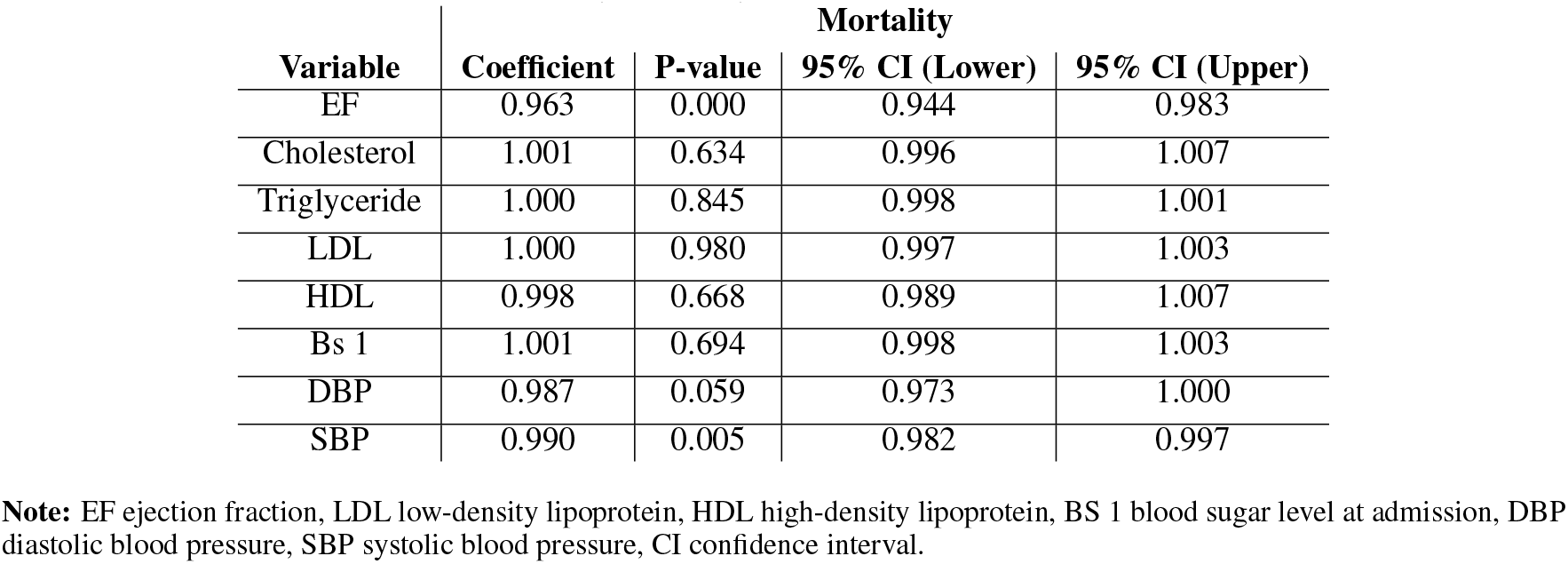
Mortality rate analysis in diabetics and non-diabetics.

**Table 5:** In-hospital mortality rate in diabetics and non-diabetics.

| Variable | Mortality |  |  |  |
| --- | --- | --- | --- | --- |
|  | Coefficient | P-value | 95% CI (Lower) | 95% CI (Upper) |
| EF | 0.971 | 0.145 | 0.933 | 1.010 |
| Cholesterol | 1.000 | 0.634 | 0.996 | 1.007 |
| Triglyceride | 1.000 | 0.823 | 0.996 | 1.004 |
| LDL | 0.999 | 0.916 | 0.988 | 1.011 |
| HDL | 0.987 | 0.599 | 0.941 | 1.036 |
| Bs 1 | 0.999 | 0.778 | 0.993 | 1.006 |
| DBP | 0.940 | 0.000 | 0.914 | 0.968 |
| SBP | 0.975 | 0.005 | 0.958 | 0.992 |

**Note:** EF ejection fraction, LDL low-density lipoprotein, HDL high-density lipoprotein, Bs 1 blood sugar level at admission, DBP diastolic blood pressure, SBP systolic blood pressure, CI confidence interval.

**Table 6:** In-hospital mortality rate in diabetics and non-diabetics.

| Variable | Mortality |  |  | P-value |
| --- | --- | --- | --- | --- |
|  | No | Yes | Total |  |
| **N** | 662 (96.5%) | 24 (3.5%) | 686 (100.0%) |  |
| **Diabetes** |  |  |  |  |
| no | 474 (72.1%) | 12 (52.2%) | 486 (71.5%) | 0.037 |
| yes | 183 (27.9%) | 11 (47.8%) | 194 (28.5%) |  |
| **HTN** |  |  |  |  |
| no | 417 (63.6%) | 12 (52.2%) | 429 (63.2%) | 0.265 |
| yes | 239 (36.4%) | 11 (47.8%) | 250 (36.8%) |  |
| **Hyperlipidemia** |  |  |  |  |
| no | 501 (76.5%) | 14 (60.9%) | 515 (76.0%) | 0.085 |
| yes | 154 (23.5%) | 9 (39.1%) | 163 (24.0%) |  |
| **smoker** |  |  |  |  |
| no | 358 (54.6%) | 19 (82.6%) | 377 (55.5%) | 0.008 |
| yes | 298 (45.4%) | 4 (17.4%) | 302 (44.5%) |  |
| **CKD** |  |  |  |  |
| no | 458 (73.4%) | 18 (75.0%) | 476 (73.5%) | 0.861 |
| yes | 166 (26.6%) | 6 (25.0%) | 172 (26.5%) |  |
| **Diabetes-HTN** |  |  |  |  |
| no | 558 (84.3%) | 16 (66.7%) | 574 (83.7%) | 0.022 |
| Yes | 104 (15.7%) | 8 (33.3%) | 112 (16.3%) |  |
| **Diabetes-lipid** |  |  |  |  |
| no | 588 (88.8%) | 17 (70.8%) | 605 (88.2%) | 0.007 |
| yes | 74 (11.2%) | 7 (29.2%) | 81 (11.8%) |  |
| **Diabetes-smk** |  |  |  |  |
| no | 602 (90.9%) | 23 (95.8%) | 625 (91.1%) | 0.408 |
| yes | 60 (9.1%) | 1 (4.2%) | 61 (8.9%) |  |
| **Diabetes-CKD** |  |  |  |  |
| no | 614 (92.7%) | 21 (87.5%) | 635 (92.6%) | 0.336 |
| yes | 48 (7.3%) | 3 (12.5%) | 51 (7.4%) |  |

**Note:** N number of patients, HTN hypertension, CKD chronic kidney disease, Diabetes-HTN combined diabetes and hypertension, Diabetes-lipid combined diabetes and hyperlipidemia, Diabetes-smk combined diabetes and smoker, Diabetes-CKD combined diabetes and CKD.

**Table 7:**
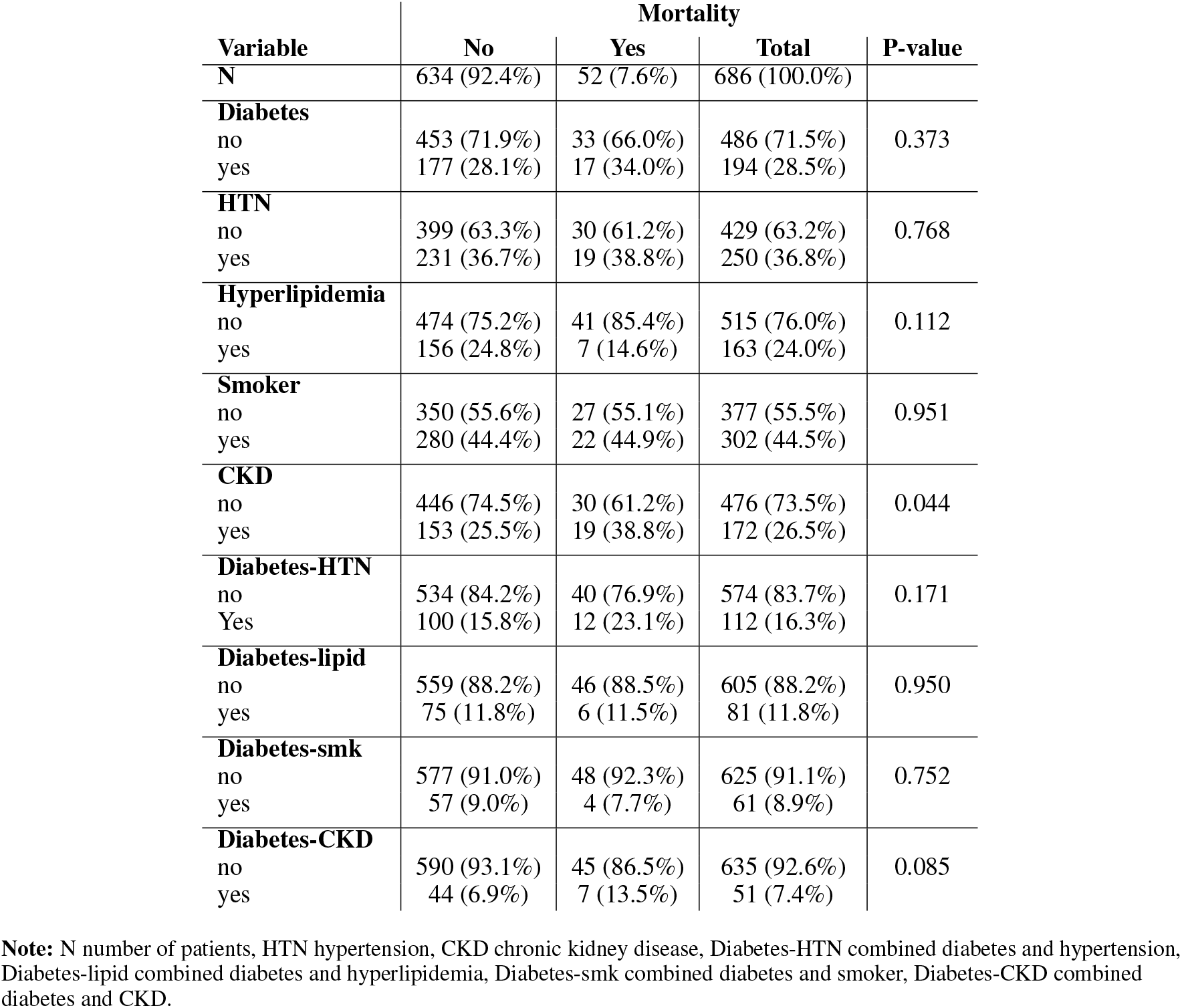
Out-of-hospital mortality in diabetics and non-diabetics.

**Table 8:** Out-of-hospital mortality rate analysis in diabetics and non-diabetics.

| Variable | Mortality |  |  |  |
| --- | --- | --- | --- | --- |
|  | Coefficient | P-value | 95% CI (Lower) | 95% CI (Upper) |
| EF | 0.960 | 0.006 | 0.933 | 0.989 |
| Cholesterol | 1.004 | 0.312 | 0.997 | 1.011 |
| Triglyceride | 0.999 | 0.522 | 0.997 | 1.002 |
| LDL | 1.001 | 0.602 | 0.998 | 1.004 |
| HDL | 1.001 | 0.872 | 0.994 | 1.007 |
| Bs 1 | 1.002 | 0.271 | 0.999 | 1.005 |
| DBP | 0.997 | 0.745 | 0.977 | 1.017 |
| SPB | 0.992 | 0.178 | 0.980 | 1.004 |

**Note:** EF ejection fraction, LDL low-density lipoprotein, HDL high-density lipoprotein, Bs 1 blood sugar level at admission, DBP diastolic blood pressure, SBP systolic blood pressure, CI confidence interval.

Lower blood glucose levels at admission were associated with an increased incidence of CVA (coefficient 0.99, *p* = 0.001). Recurrent MI was more prevalent in patients with lower ejection fraction (coefficient 0.97, *p* = 0.023). MACE was positively associated with age (coefficient 1.021, *p* < 0.001), history of MI (coefficient 1.5, *p* = 0.01), and history of CVA (coefficient 2.1, *p* = 0.017). Conversely, higher ejection fraction (coefficient < 1, *p* = 0.01) and HDL levels (coefficient < 1, *p* = 0.044) were associated with a reduced risk of MACE.

**Table 9:** CVA and intra cranial hemorrhage rate following PPCI in diabetics and non-diabetics.

| Variable | CVA and ICH |  |  |  |
| --- | --- | --- | --- | --- |
|  | Coefficient | P-value | 95% CI (Lower) | 95% CI (Upper) |
| EF | 1.029 | 0.288 | 0.976 | 1.084 |
| Cholesterol | 0.991 | 0.054 | 0.983 | 1.000 |
| Triglyceride | 1.000 | 0.818 | 0.996 | 1.003 |
| LDL | 0.993 | 0.330 | 0.980 | 1.007 |
| HDL | 1.000 | 0.973 | 0.990 | 1.009 |
| Bs 1 | 0.993 | 0.003 | 0.988 | 0.998 |
| DBP | 1.012 | 0.272 | 0.991 | 1.033 |
| SBP | 1.011 | 0.107 | 0.998 | 1.025 |

**Note:** EF ejection fraction, LDL low-density lipoprotein, HDL high-density lipoprotein, Bs 1 blood sugar level at admission, DBP diastolic blood pressure, SBP systolic blood pressure, ICH Intra Cranial Hemorrhage, CI confidence interval.

**Table 10:** CVA and intra cranial hemorrhage rate following PPCI in diabetics and non-diabetics.

| Variable | CVA and ICH |  |  | P-value |
| --- | --- | --- | --- | --- |
|  | No | Yes | Total |  |
| **N** | 544 (96.1%) | 22 (3.9%) | 566 (100.0%) |  |
| **Diabetes** |  |  |  |  |
| no | 380 (70.5%) | 14 (66.7%) | 394 (70.4%) | 0.706 |
| yes | 159 (29.5%) | 7 (33.3%) | 166 (29.6%) |  |
| **HTN** |  |  |  |  |
| no | 334 (62.1%) | 12 (57.1%) | 346 (61.9%) | 0.648 |
| yes | 204 (37.9%) | 9 (42.9%) | 213 (38.1%) |  |
| **Hyperlipidemia** |  |  |  |  |
| no | 413 (76.8%) | 17 (81.0%) | 430 (76.9%) | 0.655 |
| yes | 125 (23.2%) | 4 (19.0%) | 129 (23.1%) |  |
| **Smoker** |  |  |  |  |
| no | 303 (56.3%) | 10 (47.6%) | 313 (56.0%) | 0.431 |
| yes | 235 (43.7%) | 11 (52.4%) | 246 (44.0%) |  |
| **CKD** |  |  |  |  |
| no | 376 (72.0%) | 14 (63.6%) | 390 (71.7%) | 0.392 |
| yes | 146 (28.0%) | 8 (36.4%) | 154 (28.3%) |  |
| **Diabetes-HTN** |  |  |  |  |
| no | 453 (83.3%) | 18 (81.8%) | 471 (83.2%) | 0.858 |
| yes | 91 (16.7%) | 4 (18.2%) | 95 (16.8%) |  |
| **Diabetes-lipid** |  |  |  |  |
| no | 480 (88.2%) | 20 (90.9%) | 500 (88.3%) | 0.702 |
| yes | 64 (11.8%) | 2 (9.1%) | 66 (11.7%) |  |
| **Diabetes-smk** |  |  |  |  |
| no | 499 (91.7%) | 19 (86.4%) | 518 (91.5%) | 0.376 |
| yes | 45 (8.3%) | 3 (13.6%) | 48 (8.5%) |  |
| **Diabetes-CKD** |  |  |  |  |
| no | 500 (91.9%) | 19 (86.4%) | 519 (91.7%) | 0.355 |
| yes | 44 (8.1%) | 3 (13.6%) | 47 (8.3%) |  |

**Note:** N number of patients, HTN hypertension, CKD chronic kidney disease, Diabetes-HTN combined diabetes and hypertension, Diabetes-lipid combined diabetes and hyperlipidemia, Diabetes-smk combined diabetes and smoker, Diabetes-CKD combined diabetes and CKD.

**Table 11:** Recurrent MI rate following PPCI in diabetics and non-diabetics.

| Variable | Recurrent MI rate |  |  | P-value |
| --- | --- | --- | --- | --- |
|  | No | Yes | Total |  |
| **N** | 508 (88.7%) | 65 (11.3%) | 573 (100.0%) |  |
| **Diabetes** |  |  |  |  |
| no | 351 (69.9%) | 48 (73.8%) | 399 (70.4%) | 0.514 |
| yes | 151 (30.1%) | 17 (26.2%) | 168 (29.6%) |  |
| **HTN** |  |  |  |  |
| no | 310 (61.9%) | 40 (61.5%) | 350 (61.8%) | 0.958 |
| yes | 191 (38.1%) | 25 (38.5%) | 216 (38.2%) |  |
| **Hyperlipidemia** |  |  |  |  |
| no | 391 (78.0%) | 45 (69.2%) | 436 (77.0%) | 0.112 |
| yes | 110 (22.0%) | 20 (30.8%) | 130 (23.0%) |  |
| **smoker** |  |  |  |  |
| no | 285 (56.9%) | 30 (46.2%) | 315 (55.7%) | 0.101 |
| yes | 216 (43.1%) | 35 (53.8%) | 251 (44.3%) |  |
| **CKD** |  |  |  |  |
| no | 356 (72.8%) | 39 (62.9%) | 395 (71.7%) | 0.103 |
| yes | 133 (27.2%) | 23 (37.1%) | 156 (28.3%) |  |
| **Diabetes-HTN** |  |  |  |  |
| no | 421 (82.9%) | 55 (84.6%) | 476 (83.1%) | 0.724 |
| yes | 87 (17.1%) | 10 (15.4%) | 97 (16.9%) |  |
| **Diabetes-lipid** |  |  |  |  |
| no | 450 (88.6%) | 56 (86.2%) | 506 (88.3%) | 0.566 |
| yes | 58 (11.4%) | 9 (13.8%) | 67 (11.7%) |  |
| **Diabetes-smk** |  |  |  |  |
| no | 466 (91.7%) | 57 (87.7%) | 523 (91.3%) | 0.277 |
| yes | 42 (8.3%) | 8 (12.3%) | 50 (8.7%) |  |
| **Diabetes-CKD** |  |  |  |  |
| no | 468 (92.1%) | 58 (89.2%) | 526 (91.8%) | 0.423 |
| yes | 40 (7.9%) | 7 (10.8%) | 47 (8.2%) |  |

**Note:** N number of patients, HTN hypertension, CKD chronic kidney disease, Diabetes-HTN combined diabetes and hypertension, Diabetes-lipid combined diabetes and hyperlipidemia, Diabetes-smk combined diabetes and smoker, Diabetes-CKD combined diabetes and CKD.

**Table 12:** Recurrent MI rate following PPCI in diabetics and non-diabetics.

| Variable | Recurrent MI |  |  |  |
| --- | --- | --- | --- | --- |
|  | Coefficient | P-value | 95% CI (Lower) | 95% CI (Upper) |
| **EF** | 0.975 | 0.023 | 0.955 | 0.997 |
| **Cholesterol** | 1.000 | 0.976 | 0.994 | 1.006 |
| **Triglyceride** | 0.998 | 0.438 | 0.994 | 1.003 |
| **LDL** | 1.000 | 0.982 | 0.997 | 1.003 |
| **HDL** | 0.996 | 0.573 | 0.984 | 1.009 |
| **Bs 1** | 0.996 | 0.073 | 0.992 | 1.000 |
| **DBP** | 1.010 | 0.229 | 0.994 | 1.026 |
| **SBP** | 1.009 | 0.080 | 0.999 | 1.018 |

**Note:** EF ejection fraction, LDL low-density lipoprotein, HDL high-density lipoprotein, Bs 1 blood sugar level at admission, DBP diastolic blood pressure, SBP systolic blood pressure, CI confidence interval.

**Table 13:**
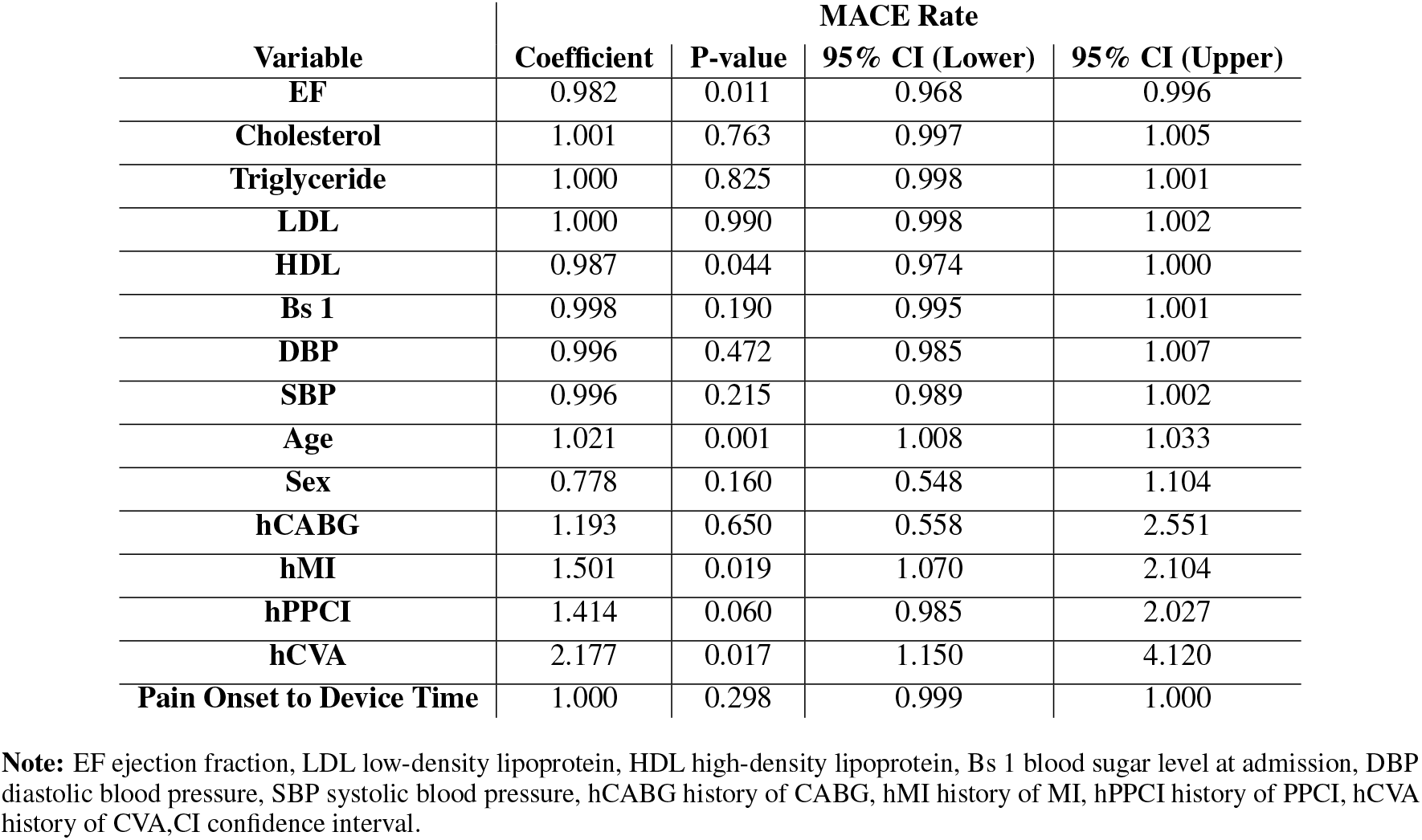
MACE incidence rate after PPCI in diabetics and non-diabetics.

**Table 14:** MACE incidence rate after PPCI in diabetics and non-diabetics.

| Variable | MACE Rate |  |  | P-value |
| --- | --- | --- | --- | --- |
|  | No | Yes | Total |  |
| **N** | 470 (76.3%) | 146 (23.7%) | 616 (100.0%) |  |
| **Diabetes** |  |  |  |  |
| no | 333 (71.2%) | 98 (69.0%) | 431 (70.7%) | 0.624 |
| yes | 135 (28.8%) | 44 (31.0%) | 179 (29.3%) |  |
| **HTN** |  |  |  |  |
| no | 298 (63.7%) | 84 (59.6%) | 382 (62.7%) | 0.377 |
| yes | 170 (36.3%) | 57 (40.4%) | 227 (37.3%) |  |
| **Hyperlipidemia** |  |  |  |  |
| no | 362 (77.4%) | 106 (75.7%) | 468 (77.0%) | 0.687 |
| yes | 106 (22.6%) | 34 (24.3%) | 140 (23.0%) |  |
| **Smoker** |  |  |  |  |
| no | 264 (56.4%) | 76 (53.9%) | 340 (55.8%) | 0.599 |
| yes | 204 (43.6%) | 65 (46.1%) | 269 (44.2%) |  |
| **CKD** |  |  |  |  |
| no | 326 (72.4%) | 99 (70.7%) | 425 (72.0%) | 0.690 |
| yes | 124 (27.6%) | 41 (29.3%) | 165 (28.0%) |  |
| **Diabetes-HTN** |  |  |  |  |
| no | 398 (84.7%) | 116 (79.5%) | 514 (83.4%) | 0.138 |
| yes | 72 (15.3%) | 30 (20.5%) | 102 (16.6%) |  |
| **Diabetes-lipid** |  |  |  |  |
| no | 414 (88.1%) | 130 (89.0%) | 544 (88.3%) | 0.753 |
| yes | 56 (11.9%) | 16 (11.0%) | 72 (11.7%) |  |
| **Diabetes-smk** |  |  |  |  |
| no | 428 (91.1%) | 132 (90.4%) | 560 (90.9%) | 0.811 |
| yes | 42 (8.9%) | 14 (9.6%) | 56 (9.1%) |  |
| **Diabetes-CKD** |  |  |  |  |
| no | 433 (92.1%) | 133 (91.1%) | 566 (91.9%) | 0.690 |
| yes | 37 (7.9%) | 13 (8.9%) | 50 (8.1%) |  |

**Note:** N number of patients, HTN hypertension, CKD chronic kidney disease, Diabetes-HTN combined diabetes and hypertension, Diabetes-lipid combined diabetes and hyperlipidemia, Diabetes-smk combined diabetes and smoker, Diabetes-CKD combined diabetes and CKD.

**Table 15:** Association between MACE and combined HTN, diabetes.

| Variable | Coefficient | SE | t-value | P-value | 95% CI (Lower) | 95% CI (Upper) | Sig |
| --- | --- | --- | --- | --- | --- | --- | --- |
| **DBP** | 1.334 | 0.391 | 0.98 | 0.326 | 0.751 | 2.370 |  |
| **Age** | 1.028 | 0.013 | 2.09 | 0.037 | 1.002 | 1.054 | ** |
| **Sex** | 0.745 | 0.200 | -1.10 | 0.273 | 0.440 | 1.261 |  |
| **hMI** | 0.536 | 0.308 | -1.08 | 0.278 | 0.173 | 1.656 |  |
| **hPPCI** | 1.317 | 0.645 | 0.56 | 0.575 | 0.504 | 3.441 |  |
| **hCVA** | 1.679 | 1.626 | 0.54 | 0.592 | 0.252 | 11.200 |  |
| **Constant** | 0.047 | 0.040 | -3.53 | 0.000 | 0.008 | 0.256 | *** |

**Note:** DBP diastolic blood pressure, hMI history of MI, hPPCI history of PPCI, hCVA history of CVA, Coef coefficient, SE standard error.

In multiple linear regression with adjusting confounding factors, there was no association between MACE and diabetes.

**Table 16:** Association between MACE and combined HTN, diabetes.

| Variable | Coef. | St.Err. | t-value | p-value | 95% Conf (Lower) | 95% Conf (Upper) | Sig |
| --- | --- | --- | --- | --- | --- | --- | --- |
| **DBP** | 1.277 | 0.29 | 1.07 | 0.282 | 0.818 | 1.993 |  |
| **Age** | 1.016 | 0.011 | 1.51 | 0.131 | 0.995 | 1.037 |  |
| **Sex** | 0.894 | 0.218 | -0.46 | 0.646 | 0.554 | 1.443 |  |
| **hMI** | 1.856 | 0.786 | 1.46 | 0.144 | 0.809 | 4.256 |  |
| **hPPCI** | 0.621 | 0.287 | -1.03 | 0.302 | 0.251 | 1.535 |  |
| **hCVA** | 2.072 | 1.069 | 1.41 | 0.158 | 0.754 | 5.694 |  |
| **Constant** | 0.083 | 0.062 | -3.36 | 0.001 | 0.02 | 0.355 | *** |

**Note:** DBP diastolic blood pressure, hMI history of MI, hPPCI history of PPCI, hCVA history of CVA, Coef coefficient, SE standard error.

In multiple linear regression with adjusting confounding factors, there was no association between MACE and hypertension.

**Table 17:** AKI rate analysis in diabetics and non-diabetics.

| Variable | AKI Rate |  |  | Test |
| --- | --- | --- | --- | --- |
|  | No | Yes | Total |  |
| **N** | 462 (80.8%) | 110 (19.2%) | 572 (100.0%) |  |
| **Diabetes** |  |  |  |  |
| No | 324 (70.9%) | 74 (67.3%) | 398 (70.2%) | 0.456 |
| Yes | 133 (29.1%) | 36 (32.7%) | 169 (29.8%) |  |
| **HTN** |  |  |  |  |
| No | 277 (60.7%) | 73 (66.4%) | 350 (61.8%) | 0.276 |
| Yes | 179 (39.3%) | 37 (33.6%) | 216 (38.2%) |  |
| **Hyperlipidemia** |  |  |  |  |
| No | 354 (77.6%) | 81 (73.6%) | 435 (76.9%) | 0.373 |
| Yes | 102 (22.4%) | 29 (26.4%) | 131 (23.1%) |  |
| **Smoker** |  |  |  |  |
| No | 260 (57.0%) | 56 (50.9%) | 316 (55.8%) | 0.247 |
| Yes | 196 (43.0%) | 54 (49.1%) | 250 (44.2%) |  |
| **CKD** |  |  |  |  |
| No | 334 (75.7%) | 60 (55.0%) | 394 (71.6%) | <0.001 |
| Yes | 107 (24.3%) | 49 (45.0%) | 156 (28.4%) |  |
| **Diabetes-HTN** |  |  |  |  |
| No | 381 (82.5%) | 95 (86.4%) | 476 (83.2%) | 0.326 |
| Yes | 81 (17.5%) | 15 (13.6%) | 96 (16.8%) |  |
| **Diabetes-lipid** |  |  |  |  |
| No | 410 (88.7%) | 94 (85.5%) | 504 (88.1%) | 0.338 |
| Yes | 52 (11.3%) | 16 (14.5%) | 68 (11.9%) |  |
| **Diabetes-smk** |  |  |  |  |
| No | 425 (92.0%) | 97 (88.2%) | 522 (91.3%) | 0.204 |
| Yes | 37 (8.0%) | 13 (11.8%) | 50 (8.7%) |  |
| **Diabetes-CKD** |  |  |  |  |
| No | 431 (93.3%) | 93 (84.5%) | 524 (91.6%) | 0.003 |
| yes | 31 (6.7%) | 17 (15.5%) | 48 (8.4%) |  |

**Note:** N number of patients, HTN hypertension, CKD chronic kidney disease, Diabetes-HTN combined diabetes and hypertension, Diabetes-lipid combined diabetes and hyperlipidemia, Diabetes-smk combined diabetes and smoker, Diabetes-CKD combined diabetes and CKD.

**Table 18:** AKI rate analysis in diabetics and non-diabetics.

| Variable | AKI |  |  |  |
| --- | --- | --- | --- | --- |
|  | Coef. | P-value | 95% CI (Lower) | 95% CI (Upper) |
| **EF** | 0.989 | 0.177 | 0.972 | 1.005 |
| **Cholesterol** | 1.001 | 0.727 | 0.997 | 1.005 |
| **Triglyceride** | 1.001 | 0.040 | 1.000 | 1.002 |
| **LDL** | 1.001 | 0.082 | 1.000 | 1.003 |
| **HDL** | 1.003 | 0.000 | 1.002 | 1.004 |
| **Bs 1** | 1.000 | 0.672 | 0.998 | 1.002 |
| **DBP** | 0.990 | 0.085 | 0.978 | 1.001 |
| **SBP** | 0.995 | 0.134 | 0.988 | 1.002 |

**Note:** EF ejection fraction, LDL low-density lipoprotein, HDL high-density lipoprotein, Bs 1 blood sugar level at admission, DBP diastolic blood pressure, SBP systolic blood pressure, CI confidence interval.

## 6 Discussion

Contrary to previous findings suggesting that diabetic patients with chronic kidney disease (CKD) experience higher mortality than those without CKD, our study did not demonstrate a significant association between the combination of diabetes and CKD and the incidence of MACE or in- and out-of-hospital mortality [17–19]. However, CKD alone was associated with increased out-of-hospital mortality. Furthermore, our data showed that acute kidney injury (AKI) was more prevalent among patients with a history of CKD, either alone or in combination with diabetes. After adjusting for confounding variables, lower triglyceride and HDL levels emerged as protective factors against AKI. Previous research has reported that diabetic patients have the highest cardiovascular mortality rates and that the coexistence of diabetes and hypercholesterolemia is a major contributor to total mortality. Consistent with these findings, our study demonstrated higher rates of prior MI, PPCI, and multivessel involvement in diabetic patients. Both smoking and diabetes were associated with elevated overall mortality. In-hospital mortality was notably higher among diabetic patients with concomitant hyperlipidemia. Additionally, hypertension combined with diabetes was associated with increased in-hospital mortality. However, no significant relationship was found between diabetes, either isolated or in combination with hypertension and the incidence of MACE, CVA, or in-hospital mortality [20]. A recent study assessing mortality in diabetic patients post-PPCI found no significant difference in mortality between diabetics treated with oral antidiabetic medications and the general population, although mortality was higher among insulin-treated patients. Our findings are in agreement, showing that both in-hospital and 30-day mortality rates were higher in diabetic patients compared to non-diabetics [21]. A previous study reported no association between diabetes and in-hospital or short-term Mortality while as there was connection between diabetes and long-term mortality. Our results did not identify a link between diabetes and out-of-hospital mortality despite the observed relationship between CKD and out-of-hospital mortality [22]. Previous literature has indicated a higher likelihood of recurrent MI and cardiovascular mortality in diabetic patients. Although our findings aligned with the increased cardiovascular mortality, we found no significant association between diabetes and recurrent MI. Moreover, in comparison with patients with isolated hypertension, diabetic Patients, either alone or in combination with hypertension, showed higher in-hospital and overall mortality. In line with previous findings, age, CKD, history of MI, and CVA were identified as significant predictors of total mortality. Both MI and CVA history were also associated with increased cardiovascular risk, further supporting our results [23]. Interestingly, some studies have reported that hyperglycemia may reduce mortality risk in diabetic patients. Similarly, our data suggest that stress hyperglycemia was associated with a lower risk of CVA in STEMI patients and did not increase the prevalence of MACE [24].

## 7 Conclusion

In patients undergoing primary PCI for STEMI, diabetes, smoking, and the presence of comorbid conditions such as hypertension and hyperlipidemia were associated with increased short- and long-term mortality. Higher ejection fraction and systolic blood pressure at admission were protective factors against in-hospital mortality, whereas the history of kidney disease significantly predicted higher long-term mortality. Additionally, cerebrovascular accidents (CVA) post-PCI were associated with lower blood sugar levels, and recurrent myocardial infarction (MI) was more frequently observed in patients with reduced ejection fraction. Major adverse cardiovascular events (MACE) were primarily influenced by age, history of MI, and history of CVA, while elevated ejection fraction and HDL levels demonstrated protective effects. These findings emphasize the complex interplay between clinical factors and medical history in shaping outcomes after primary PCI.

## Data Availability

All data produced in the present work are contained in the manuscript.

